# The efficacy of candidate HIV vaccines in non-human primates and humans: a systematic review

**DOI:** 10.1101/2025.11.21.25340585

**Authors:** Julia M. L. Menon, Marianna Rosso, Marjolein van de Weerthof, Frederique Struijs, Astrid van Hattem, Larissa Prechal, Olivier Weber, Benjamin Victor Ineichen, Cathalijn H. C. Leenaars, Merel Ritskes-Hoitinga

**Affiliations:** Netherlands Heart Institute, Moreelsepark 1, 3511 EP Utrecht, Netherlands; Department Population Health Sciences, Institute for Risk Assessment Sciences Toxicology (IRAStox), University Utrecht, Yalelaan 104-106, 3584CM Utrecht, The Netherlands; Center for Reproducible Science, University of Zurich, Zurich, Switzerland; Department of Clinical Research, University of Bern, Bern, Switzerland; Institute for laboratory animal science, Hannover Medical School, Hannover, Germany; Department of Clinical Medicine, Aarhus University, Aarhus University Hospital, A1001, Palle Juul-Jensens Boulevard 99, 8200 Aarhus N, Aarhus, Denmark

**Keywords:** Systematic review, meta-analysis, efficacy, HIV, vaccines, non-human primates

## Abstract

Over the last four decades, Acquired Immunodeficiency Syndrome (AIDS) and Human Immunodeficiency Virus (HIV) have been studied intensively in animal models and humans, hoping to find an effective and safe vaccine to prevent this debilitating disease. However, no vaccine has become available, and the translatability of the animal models to humans has not been assessed systematically. Our systematic and comprehensive review of published HIV trials in non-human primates (NHPs) and humans aimed to answer the question: “What is the concordance of candidate HIV vaccine efficacy in NHPs and humans?”.

A review protocol was posted on Prospero on 20 December 2023. We searched PubMed and EmBase on 25 and 26 December 2023 with comprehensive search strings, and used a combination of AI-assisted and manual screening (two independent screeners). Data extraction followed a custom-made template, and risk of bias was evaluated following standard tools (both by two independent reviewers).

From 50 624 records retrieved by the searches, 216 were included, comprising data from 40 750 human subjects and 3 326 NHPs. Differences between NHP and human studies were observed for e.g. the number and type of vaccines administered, the control conditions and the level of infection, further discussed in the manuscript. Our explorative frequentist random-effects model meta-analysis shows that overall, the NHP and human results were not significantly different (Q=1.75; df=1; p=0.19). However, risk of bias was mainly unclear and confidence intervals were large.

To conclude, limited animal-to-human translation cannot be held responsible for the current lack of an effective preventive HIV vaccine, but the results do not support the value of NHP models either. All extracted data are available from a searchable online spreadsheet, to aid those working on HIV vaccine development in more easily retrieving relevant studies.

## Introduction

Over the last four decades, many scientists have dedicated substantial time and resources studying Acquired Immunodeficiency Syndrome (AIDS). The first scientific publication of this debilitating and deadly disease dates from 1981 (1). The first major breakthrough in AIDS research was isolation of the Human Immunodeficiency Virus (HIV) (2), which was followed by many important milestones, comprising the elucidation of HIVs structure, sequence, replication mechanism and pathology; the identification of different groups and clades; and the development of diagnostic testing, effective antiretroviral treatment, and prophylactic measures (3–7). While these successes may seem reason for optimism, numbers of newly infected individuals are expected to keep increasing (5, 8) and treatment is far from accessible to all. Besides, lifelong antiretroviral treatment is associated with comorbidities and adverse events in people living with HIV (PLWH), which increase over time (7, 9). Thus, HIV remains a global health concern for which an effective and safe vaccine is needed.

Several models have shown that, depending mainly on efficacy, price, and HIV incidence in the target population, HIV vaccines would be a cost-effective healthcare measure (10). Various types of vaccines were sequentially developed, and many of them even tested in clinical trials. The tested classical and more modern approaches comprise inactivated and live-attenuated virus, subunit, vectored and DNA vaccines. These approaches aim for induction of neutralizing antibodies, cell-mediated immunity, boosting the innate immune system, and combinations of those immune responses (3, 6, 11, 12). However, at the time of submission of this manuscript, no preventive HIV vaccine is available, nor does any seem to have advanced far enough through the pharmaceutical pipeline to becoming available anytime soon.

There are many challenges in any kind of pharmaceutical development, from creating an efficacious and safe formulation, through regulation and cost-benefit balance to public acceptance (13, 14). Vaccine development may well be more challenging than development of other drugs, both because of the complexity of the immune response, and because of vaccines generally being administered to large numbers of healthy individuals, which necessitates high safety levels. In the case of HIV preventive vaccines, development is further complicated by the virus’ effective ability to evade the immune system in six different ways. These are, first, disguising itself by incorporating host-antigens in its envelope; second, rapidly mutating surface antigens; third, only very briefly and sterically shielded exposing of conserved viral regions; fourth, some tolerance to variation even in the conserved regions; fifth, latently infecting CD4+ cells acting as reservoirs; and sixth, down-regulating the immune cells that are needed to create an effective immune response (15–18).

The fickly nature of HIV is clear from the near-absolute lack of spontaneous recovery and survival of infection (17, 19), which is in clear contrast to most other viruses for which vaccines have been developed. A further challenging difference between HIV and other infectious diseases, for which vaccines *are* available, is that so far, elicited antibody responses are much shorter, with half-lives of 8 weeks, compared to documented serum presence up to 26 years (with half-life estimates of 50-200 years)(16, 20). Most failures of early HIV-vaccine candidate clinical trials may now be explained by the wealth of mechanistic knowledge becoming available since, but over four decades of research have not yet established a reproducible correlate of protection (CoP) to work with (12, 18, 21–23). Thus, future development of an effective HIV-vaccine is still far from certain (12, 24–26).

Non-human primates (NHPs) have been a preferred animal model in preclinical HIV vaccine research and development. However, poor animal-to-human translatability could hinder research progress (17, 27). The general challenge of animal-to-human translation has been a topic of discussion within the laboratory animal science field, but most discussions on the predictability of animal models for human healthcare are based on qualitative comparisons that are not very reliable(28). Five quantitative analyses of animal-to-human translation shed some light on the predictive validity of animal models for human health care. First, Lindl et al. followed up the results from 51 animal ethics requests, and found very little evidence of translation to the clinical situation (29). Second, Hackam et al. followed up on 76 highly cited animal studies, and found that about one third had contributed to later randomized clinical trials (30). Third, Contopoulos-Ioannidis et al. followed up on 101 articles that described novel therapeutic or preventive promises based on animal data (31). Only 16 of these were tested in clinical trials, of which 12 had a positive outcome. Fourth, Perel et al. compared the effects of 6 interventions between animals and humans with systematic literature reviews per intervention, with half of them concordant (32). Fifth, a systematic scoping review of reviews showed that translational success rates ranged from 0 to 100% (28). In that scoping review, individual factors could not predict successful translation, but combinations of specific analysis design factors might. Further analyses of those data showed that the combination of recent small reviews with analyses at the event or study (but not the intervention) level, including more than one species, and using a quantitative definition of translation, seemed more consistent with successful translation than other combinations (33).

Theoretically complicating animal-to-human translation of vaccines, interspecies variability of immune systems may well be larger than variation in systems targeted by other pharmaceuticals. However, the overall animal-to-human translational success rate for the field of infectious and parasitic diseases is within the same range as for other medical fields (34). Moreover, a large pharmaceutical development database analysis showed that the probability of advancing through the development pipeline was similar for prophylactic vaccines and other pharmaceuticals. Preclinical development lasted a good year longer on average for prophylactic vaccines than for other pharmaceuticals, but it was equivalent for the subsequent three human clinical trial phases (35). Nevertheless, preclinical HIV research is hindered by an additional challenge; HIV does not infect any of the readily available animal models (6, 27). This means that animal models either have to be humanized, or that vaccines should be targeted and responses should be measured for similar viruses such as Simian Immunodeficiency Virus (SIV) or Simian HIV (SHIV), which may limit the external validity of the model.

While human HIV-infection and NHP SIV-infection share many similarities, comprising viral replication patterns, immune evasion strategies and pathological features, relevant differences in disease progression and vaccine responsiveness are present. Moreover, NHP models generally use challenges that are considered to be more aggressive than natural human viral exposure (6, 12, 19, 23). Thus, limited efficacy results in NHP trials have been followed by comparable human trials, hoping for better outcomes with natural exposure. While ideas on how to improve animal-to-human predictability specifically for vaccine efficacy have been reviewed before (36), the concordance of outcomes between NHP and human HIV trials had not yet been assessed systematically.

In this systematic review (SR) the author team comprehensively collected all published findings about HIV trials in NHPs and humans, and systematically extracted and summarized the findings to assess concordance of the trial designs and results between species. Our main review question was: “What is the concordance of candidate HIV vaccine efficacy in non-human primates and humans?”, which we answered by assessing the efficacy of candidate HIV vaccines compared to non-vaccinated controls in both NHPs and humans, and pooling the results in a single translational meta-analysis.

## Methods

A protocol for this review was posted on Prospero on 20 December 2023 (registration number: <u>CRD42023495529</u>). In this manuscript, we provide a summary and operationalizations of our definitions. Minimal protocol deviations are described at the end of the methods section. In this publication, we perceptively followed the PRISMA guidelines (37).

## Search

PubMed (https://pubmed.ncbi.nlm.nih.gov/) was searched on 25 December 2023 and EMBASE via Ovid on 26 December 2023. The search strategy was based on three search components: the population (“non-human primates”, using established search filters from Cassidy et al. (38) and “humans”, using a previously developed search string from Van de Wall et al. (34)), the intervention (“vaccines”) and the field of interest (“HIV”). For each component we used a combination of MeSH/Emtree terms besides title-abstract-keyword terms, the latter including variations in spelling and plurals. No publication date or language restrictions were applied. The full search strategy is provided both in the supplementary data for this manuscript on the Open Science Framework (OSF, https://osf.io/zkmcp) and as an appendix (Appendix A: search criteria – full list of search terms).

To facilitate exports of the large numbers of records, and in line with later screening in batches (described below), separate searches were performed for NHP and human studies on the same date (per database; two sequential days for the two databases). The studies retrieved by both searches were taken out of the “human” search results via “Boolean-NOT” searches, to prevent unnecessary screening of the same studies twice. The distinct searches resulted in a single evidence-stream; any type of study could be included in any batch, and batches were treated similarly except for screening (described below). However, the split was made based on the presence of NHP-related terms in title, abstract and keywords, thus the first batch contained most NHP-related papers, and the second batch most human studies. Duplicates between databases were removed with http://dedupendnote.nl/.

## Screening

All screening followed the SR-standard two-step approach; a preselection in a title-abstract-keyword phase, followed by a full text phase. The inclusion and exclusion criteria for both phases are shown in Table 1, with our operationalizations. As screening was split over two batches, there were four screening phases: title-abstract-keyword and full-text screening for the first and second batch. For the first batch, comprising all results retrieved with the NHP-specific part of the search, all screening was performed manually in Rayyan (https://rayyan.ai/). Discrepancies were resolved by discussion between screeners. Manually screening the NHP-part of the search had the following advantages: it prevented missing publications on rare NHP species, it made sure we manually checked most of the more variably designed animal studies, and it created a reference set to use for simulation studies to determine the optimal settings for AI-assisted screening.

**Table 1:**
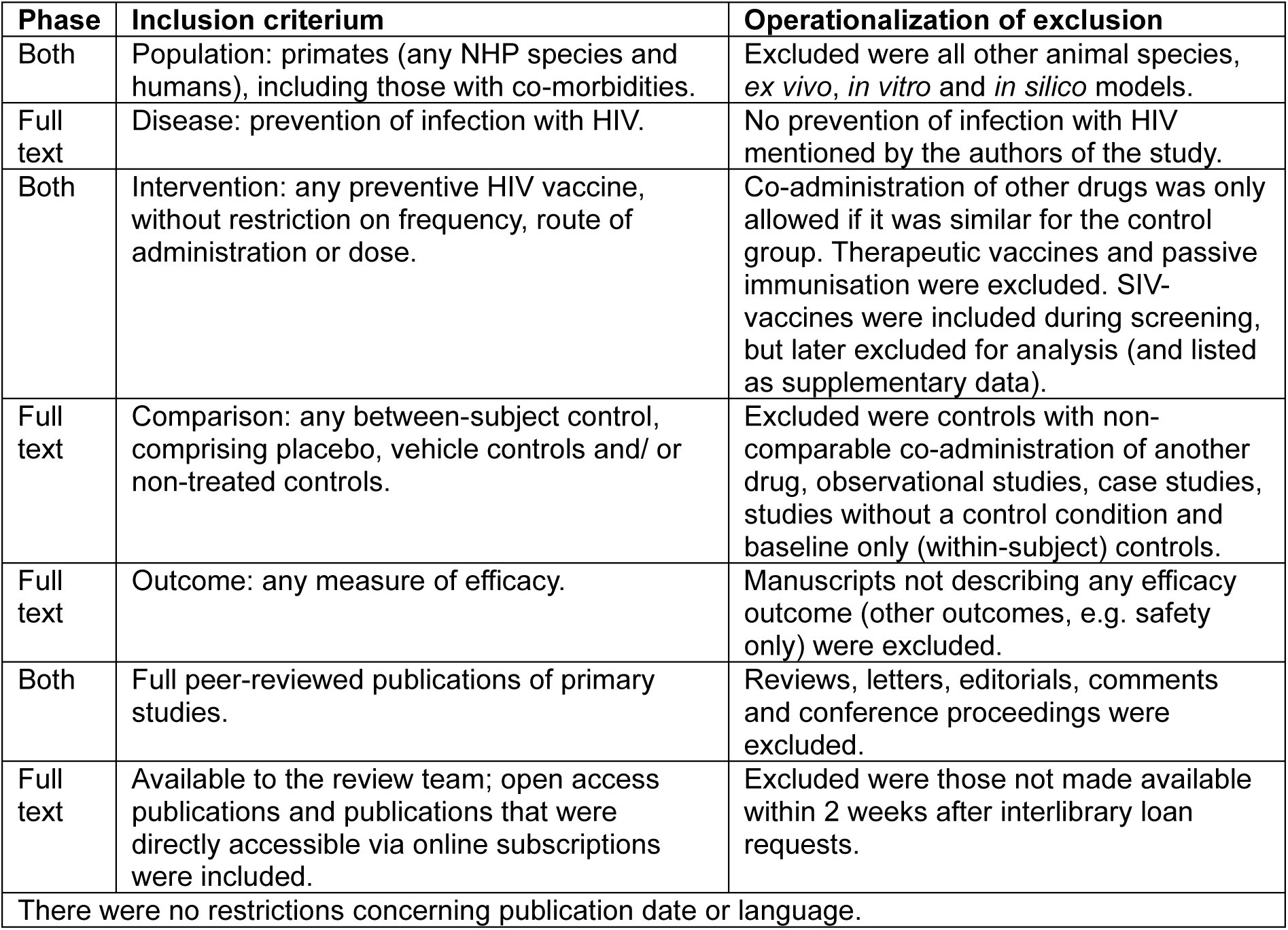
Inclusion and exclusion criteria (ranked in order)

For the second batch, title-abstract screening was performed using priority screening in ASReview (https://asreview.nl/). Screening of the second batch was largely in line with the previously described SAFE procedure (39). There were two deviations from that procedure. First, we did not identify and work with any key papers upfront, to prevent bias. Second, we adapted the procedure to work with two independent screeners. According to SAFE, different models should be used sequentially by one screener, while our two screeners used them in parallel. One screener used the text frequency – inverse document frequency feature extractor with logistic regression modelling, the other screener used the document-to-vector feature extractor with random forest modelling. Each screener used a different set of five included and five excluded records as prior information.

Model selection, settings and the stopping rule were optimized based on simulation studies, which were performed before the start of batch two screening, and described fully in a supplementary file (https://osf.io/zkmcp). The stopping rule was similar for both screeners and two-fold; first, no extra included records were identified in the last 200 screened records, and second, either 40% of the records had to be seen (i.e. 10 325), or no extra included records had been identified in the last 800 records, based on the 32% inclusion rate in the screening of batch 1 with expected differences in inclusion rate between batches. The AI never decided which studies were included and excluded, it only prioritized the studies in order of expected relevance, which was updated with every decision. Discrepancies, i.e. records identified by only one of the AI-assisted screeners, were resolved using Rayyan.

Reference lists of included studies and relevant reviews were manually screened for additional publications meeting our inclusion criteria. The reference flow of studies throughout the screening, showing how the inclusion and exclusion of reference were made, is illustrated by a flow chart shown in the result section (Figure 1).

**Figure 1:**
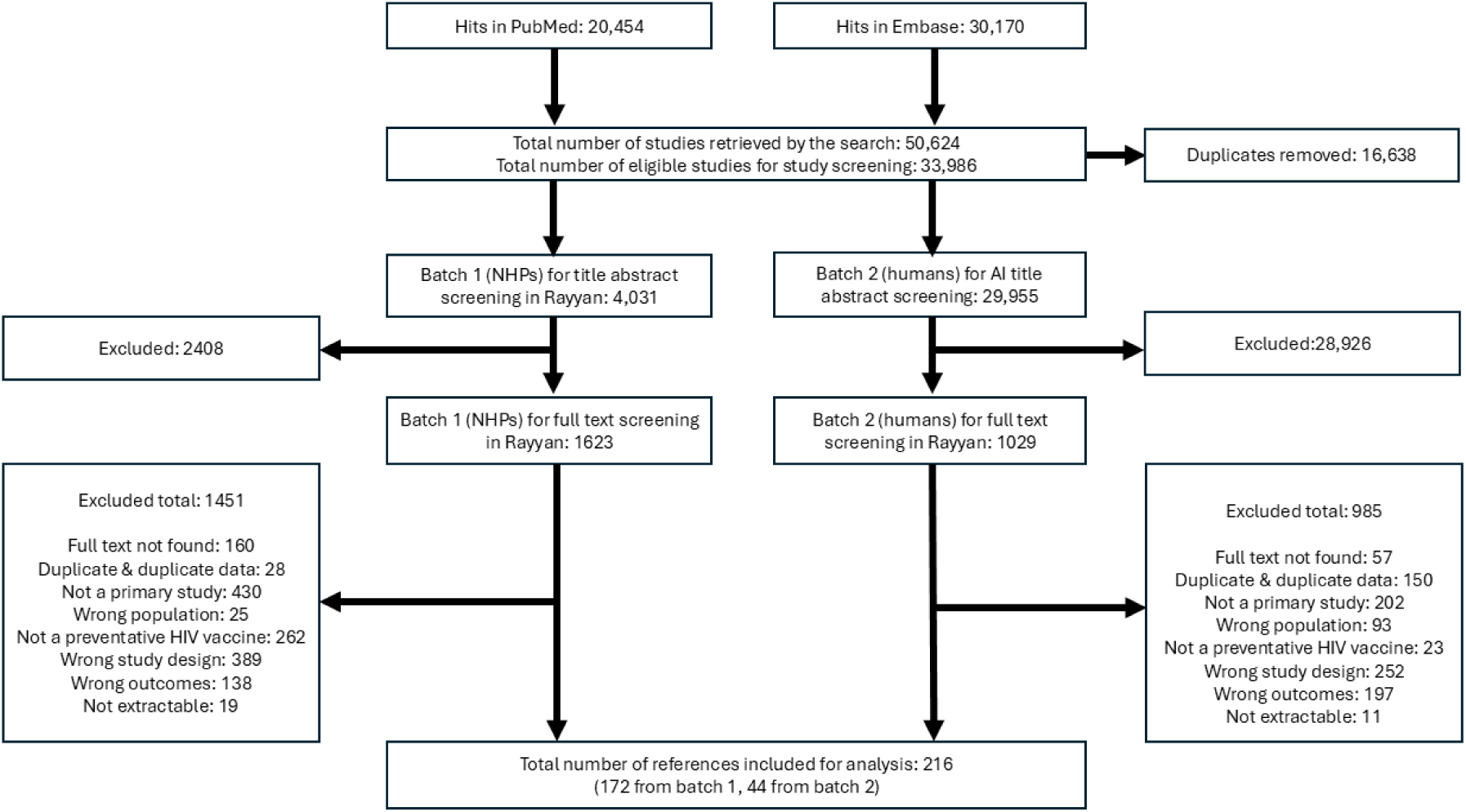
Reference flow throughout the screening process (flow chart)

## Data extraction

Data extraction was performed by two independent reviewers using a custom-made template in Covidence (https://get.covidence.org/). Data were extracted from text, tables and graphs. For graphs, we used GIMP as described previously (40) and WebPlotDigitizer (41) for graphs with logarithmic scales. Discrepancies were resolved by discussion between extractors. The data extraction comprised: 1. bibliographic data (Author, year, title, journal, pages, DOI, language, country of ethical review), 2. study design data (number of individuals/animals per group, study duration, type of control), 3. Population data (for animal studies: species, sex, body weight, age, methods of HIV infection, co-morbidities, for human studies: gender, age, exposure risk, co-morbidities), 4. intervention data (vaccine name as provided by the authors, vaccine category, formulation, dose, schedule, boosters, route of administration), 5. outcome data (author-reported efficacy measure, observed number of cases per study group for vaccine and control, risk ratio, risk difference, other measures of efficacy) and 6. Risk of bias data (see below). The full data extraction sheet, with extracted data, can be found on the Open Science Framework (OSF, https://osf.io/zkmcp) under “Extracted Data”.

For most analyses, the unit of analysis was the included paper. Scientists often combine different interventions in a single study, and multiple experiments in a single paper, posing challenges in data management for reviewers. For this review, if a paper described multiple HIV vaccination schedules meeting the inclusion criteria in separate subjects, we extracted data for all groups meeting the inclusion criteria separately. Only data from the most comparable control group(s) were extracted. To prevent the same subjects contributing multiple data points into the analyses, which would have caused issues with data dependency, we extracted a single time point when multiple time points were provided. The selected time point was generally the last time point within the study period. The only exception was for measurements of viral presence in NHP studies, because, when a substantial part of the NHPs die during the study period, analysing the last time point would have decreased the overall power of our analyses. Thus, for those studies, we extracted and analysed the latest time point where more than 50% of the sample was still alive.

Units (e.g. weeks and days for time) and measures of variation (SEM, SD) were harmonised during data extraction.

## Risk of Bias

Two independent reviewers performed risk of bias assessments for each included reference, using a custom-made template based on the relevant questions from both SYRCLE’s risk of bias tool (42) and the Cochrane risk of bias tool (43). Discrepancies were resolved by discussion or with the help of a 3rd reviewer. Compared to the SYRCLE tool, our evaluation of selective outcome reporting was more stringent; a low risk of bias score for this type of bias would only have been given if a protocol of the experiment was published upfront. We focused our risk of bias evaluations on four main study quality elements: randomization, blinding, study sample size and conflicts of interest. The evaluated elements are summarized in Table 2.

**Table 2:** Evaluated risk of bias elements.

| **Bias Element** | **Signalling question** | **Risk of bias scores** |
| --- | --- | --- |
| Selection bias - sequence generation | Was the allocation sequence adequately generated and applied? | High |
|  |  | Low |
|  |  | Unsure |
| Selection bias - baseline characteristics | Were the groups similar at baseline or were they adjusted for confounders in the analysis? | High |
|  |  | Low |
|  |  | Unsure |
| Selection bias - allocation concealment | Was the allocation adequately concealed? | High |
|  |  | Low |
|  |  | Unsure |
| Performance bias - random housing* | Were the animals randomly housed during the experiment? | High |
|  |  | Low |
|  |  | Unsure |
|  |  | Not Relevant (human study) |
| Performance bias - blinding | Were the caregivers and/or investigators blinded from knowledge which intervention each subject received during the experiment? | High |
|  |  | Low |
|  |  | Unsure |
| Detection bias - random outcome assessment | Were animals / participants selected at random for outcome assessment? | High |
|  |  | Low |
|  |  | Unsure |
| Detection bias - blinding of outcome assessment | Was the outcome assessor blinded? | High |
|  |  | Low |
|  |  | Unsure |
| Attrition bias - incomplete outcome data | Were incomplete outcome data adequately addressed? | High |
|  |  | Low |
|  |  | Unsure |
| Reporting bias - selective outcome reporting | Are reports of the study free of selective outcome reporting? | High |
|  |  | Low |
|  |  | Unsure |
| Other sources of bias | Was the study apparently free of other problems that could result in high risk of bias? | High |
|  |  | Low |
|  |  | Unsure |

## Analyses

Quantitative summary values were calculated and figures were created in R (44) via RStudio (45), using the following packages: tidyr, dplyr, stringr, janitor, tibble, ggplot2, ggpubr, readr, readxl, kableExtra, scales, tm, wordcloud, and meta (46–59). For comparisons of study characteristics between NHP and human studies we mostly created plots without performing statistical analyses (minimizing the risk of false-positive findings while creating insight into the data distributions). The only included paper containing both NHP and human data was filtered out for some of these comparisons because keeping it in would have required extensive changes in the data management process while the added informative value would have been minimal.

According to our protocol, we performed a frequentist random effects meta-analysis of overall efficacy, using a random effects model of relative risks, pooling results for studies reporting numbers infected/ diseased / dead. I^2^ was used as a measure of heterogeneity. To prevent the same subjects contributing multiple data points into the analyses, which would have caused issues with data dependency, we only included a single outcome per experimental group into these analyses. For this, we used the following hierarchy: 1. number / percentage infected, 2. presence of virus in tissue/ blood, 3. survival/ death. Subgroup analyses were performed for species (NHP versus human), but not for categorised study duration (time-at-risk in days) or categorised exposure (dose), as the study designs were too variable to create reliable categorisations. For the same reason, no sensitivity analyses were performed. A funnel plot and trim and fill analysis were performed to assess potential small study effects.

## Protocol deviations

Our main protocol deviation is not contacting authors of included studies for missing data, because we retrieved and included more references than anticipated, and none of the included studies reported all data of interest. Thus, the amount of work required to gather the missing information was not viable within the timeframe of the review. A smaller protocol deviation was that data cleaning was not performed in a spreadsheet as planned, but either in Covidence or directly in R. The reason for this was a redistribution of tasks among the review team members. The third, also small, protocol deviation was that we did not extract data on NHP strains, because we could not reliably do so. The last protocol deviation is that we decided against performing subgroup analyses for categorised study duration (time-at-risk in person-months/ animal-months) and categorised vaccine exposure (∼dose), as we could not reliably categorise those variables in a meaningful manner.

While not strictly protocol deviations, we had to refine and restrict some operationalisations of definitions throughout. We started with wider definitions and operationalisations because we were hoping to provide a wider overview than here-presented, e.g. of different proxy-outcomes for efficacy. Where adaptations were made, the records evaluated before the adaptation were re-evaluated and adapted for consistency. This manuscript clearly describes the final operationalisations.

## Results

The PubMed search retrieved 20 454 records and the EmBase search 30 170. From the combined total of 50 624 records, 16 638 duplicates were removed, resulting in 33986 potentially relevant records for screening. Of these, 4 031 (batch 1, mainly NHP studies) were imported into Rayyan for manual screening, the remaining 29 955 (batch 2, mainly human studies) were screened with AI-assistance using ASReview. During title-abstract screening, (2 408 + 28 926 =) 31 334 papers were excluded, resulting in (1 623 + 1 029 =) 2 652 papers for full text screening. Of those, (1 451 + 985 =) 2 436 were excluded, resulting in (172 + 44 =) 216 papers included in this review. Paper flow throughout the screening process is shown in Figure 1, and references in each stage of the process are available on OSF (https://osf.io/zkmcp). NHP studies testing SIV-vaccines as a model for HIV vaccines (k=184) were included during screening as we originally hoped to analyze them as a separate group of potentially relevant NHP model studies, but later excluded for analysis due to time restrictions. A list of these references is also provided on OSF (https://osf.io/zkmcp).

The included references were published from 1991 through 2023 (median: 2007) and all were written in English. According to the place of ethical review, most included studies were performed in the US (k=78), the only other country with more than 10 included studies was Japan (k=12). The three most commonly included journals were the Journal of Virology (k=41), Vaccine (k=28) and Virology (k=25). Other journals were included fewer than 10 times. The included papers described studies in humans (k=46, n= 40 750) and NHPs (k=170, n= 3 326), both categories including a single paper (60) describing results from both humans (n=36) and NHPs (n=30). NHPs were Macaques (k=167), Chimpanzees (k=2) and Baboons (k=1). Most human studies (k=43) included male and female subjects. In NHP studies, it was common to restrict testing to one of the sexes with k=24 exclusively male NHP studies and k=24 exclusively female NHP studies next to k=21 NHP studies of both sexes. In 102 NHP studies, the sex of the animals was not specified.

Early publications mainly focused on classical vaccination strategies (inactivated and live-attenuated virus), over time we see more variation in vaccination strategies, with modern vaccines (mainly DNA, protein-based, vector-based) and combinations of different strategies (Figure 2). We also observe that over time, the number of publications increases (a pattern commonly observed in literature review), but after a peak the numbers decrease again, possibly reflecting a decline in research funding (61).

**Figure 2:**
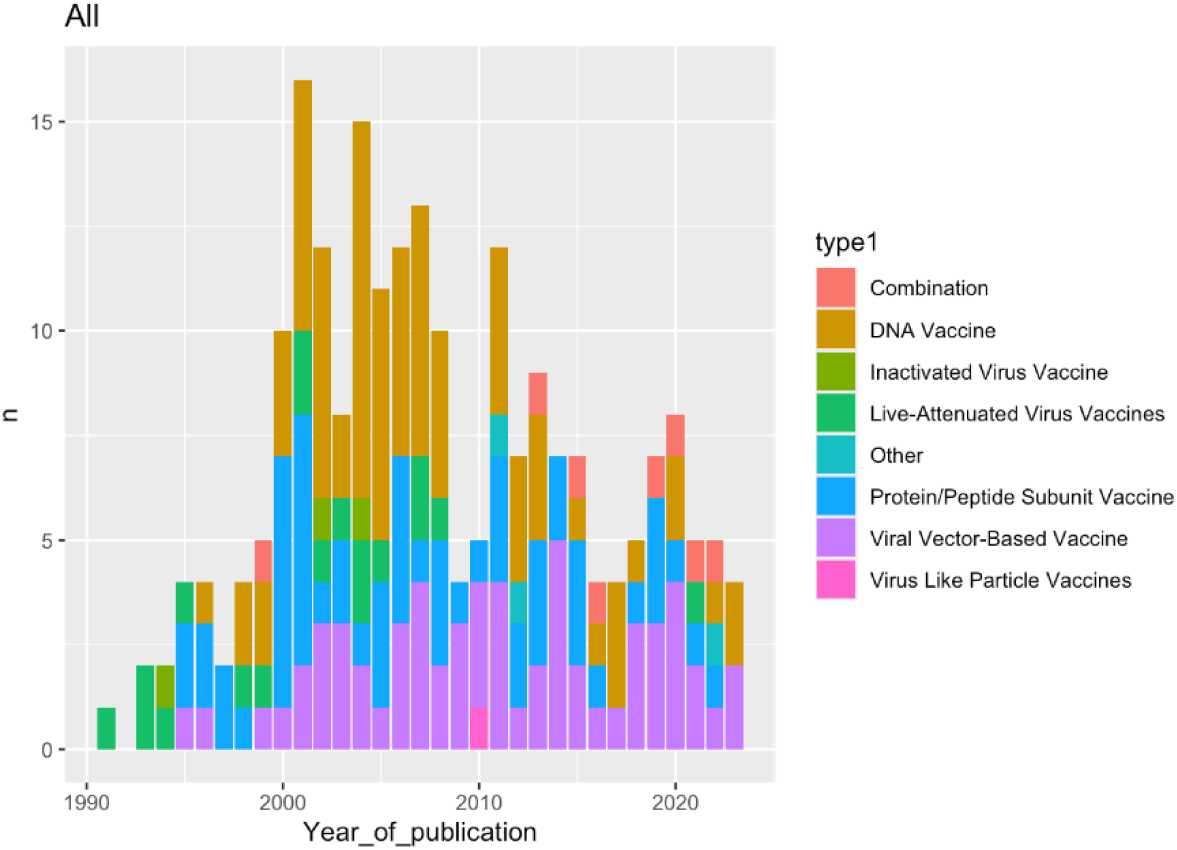
Included publications by vaccine type over time.

We did not perform statistical analyses comparing bibliographic data or study characteristics between NHP and human studies, but exploratively analyzed them with tabulations and data visualizations. As expected, on average animal studies (published from 1991 through 2023, median: 2005) seem to precede human studies (published from 1996 through 2023, median: 2012, Figure 3).

**Figure 3:**
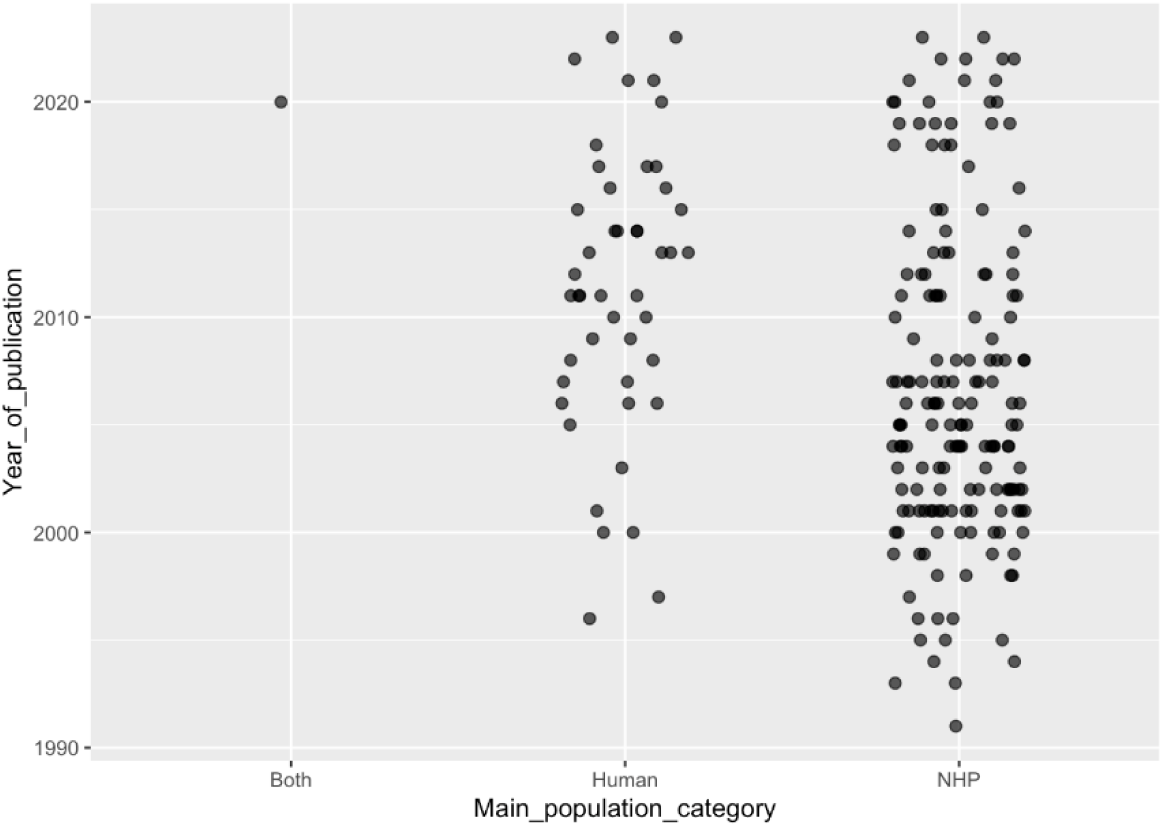
Publications dates of included NHP and human studies.

The number of tested subjects per group ranged from 1 to 8198. For NHP studies, the range was from 1 to 31 (median: 5); for human studies from 2 to 8198 (median: 17, Figure 4).

**Figure 4:**
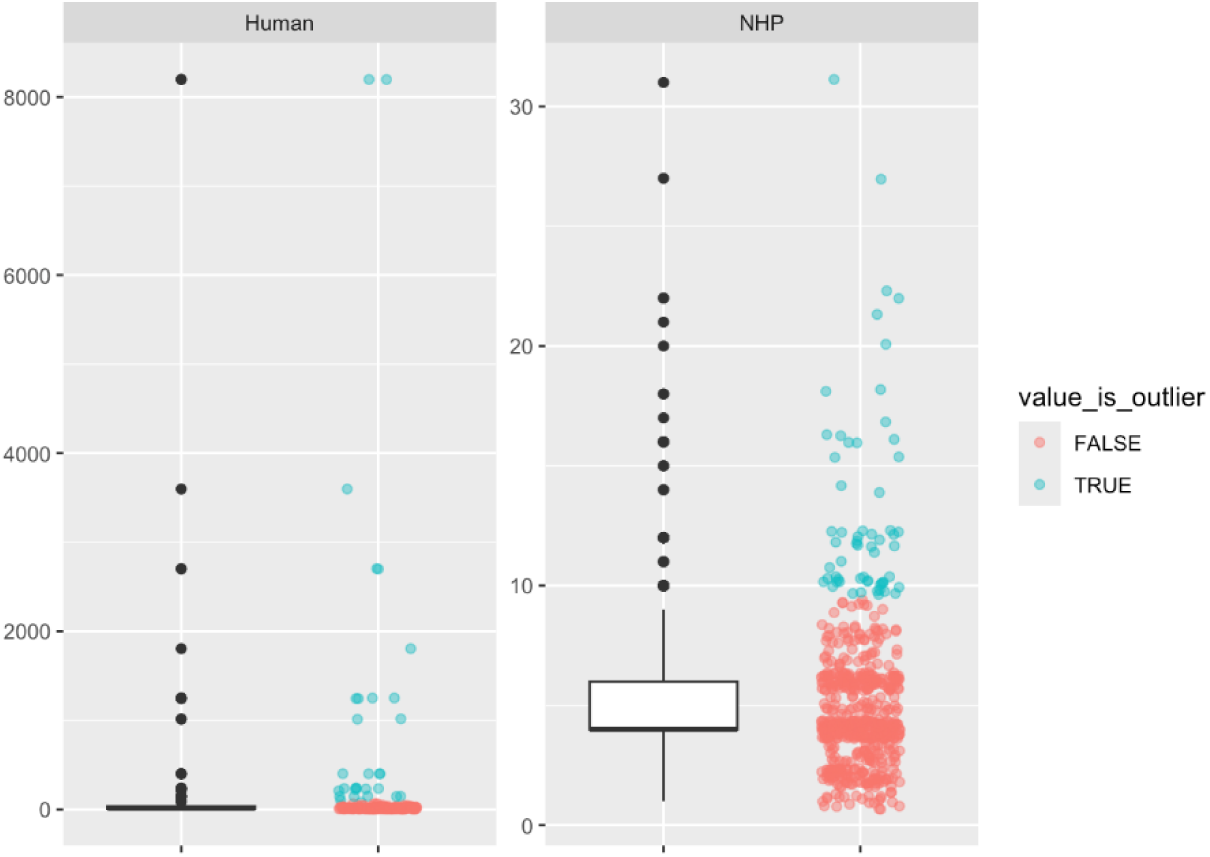
Numbers of included subjects per experimental group in NHP and human studies. In each panel, the left part shows a classical box-plot with whiskers, while the right part shows the separate values of the included study groups (with a horizontal scatter to improve visibility of overlapping values).

All included human studies tested an HIV vaccine. Most of the included NHP studies tested a SHIV vaccine (k=92), but HIV vaccines were also common (k=83, included in both counts are 5 studies testing both). The most common vaccine types were Viral Vector-Based (k=24); DNA (k=13); and Protein/Peptide Subunits (k=10) for human studies, and DNA (k=60); Protein/Peptide Subunit (k=47); Viral Vector-Based (k=38); and Live-Attenuated Virus (k=18) for NHPs. We extracted author-provided vaccine names. Terms related to the targeted epitopes and mentioned at least ten times in those names were: env (k=50), gag (k=36), tat (k=15), gp120 (k=14), gp140 (k=13), sivmac239 (k=12), gp160 (k=11) and 89.6p (k=10). Note that these numbers are underestimates because we did not correct for typographical errors and variant spellings. The most common route of administration was intramuscularly, both in humans and in NHPs. Other administration routes mentioned in more than five papers were Intravenous, Intranasal, Intradermal and Subcutaneous.

The most commonly tested adjuvants were Alum (k=13), MF59 (k=10), incomplete Freund’s adjuvant (k=6), the purified *Quillaja saponaria* extract QS-21 (k=5), Bupivacaine (k=4) and CFA (k=4). These adjuvants have been combined with different types of vaccines in both NHPs and humans as shown in Figure 5.

**Figure 5:**
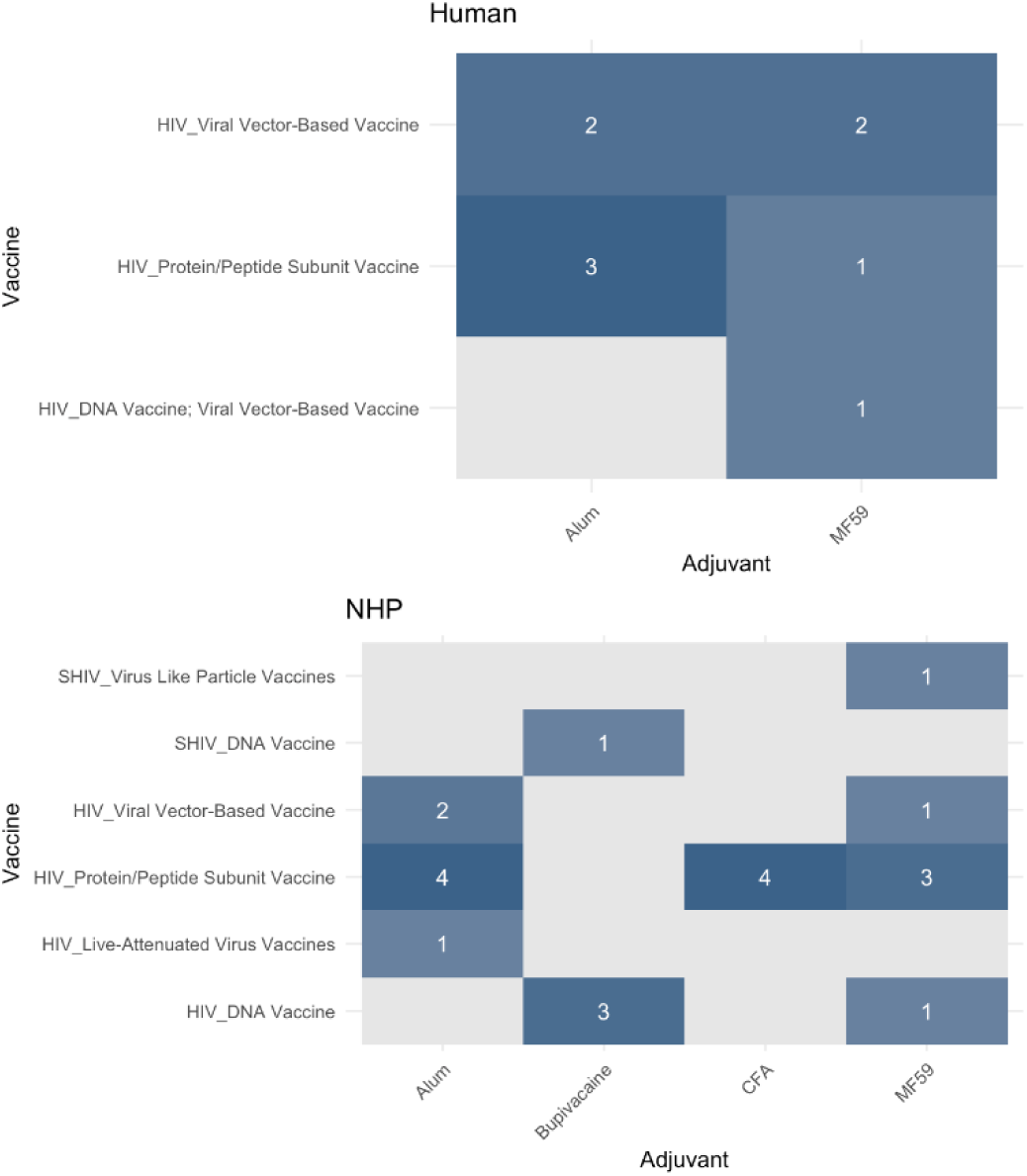
Heatmap of vaccine-adjuvant combinations in humans and NHPs.

The number of administered vaccine doses ranged from 1 to 14 for NHP studies (Median: 4), and from 1 to 7 (median: 3.5) for human studies (Figure 6). Different vaccine – booster combinations have been tested in both NHPs and humans, as shown in Figure 7.

**Figure 6:**
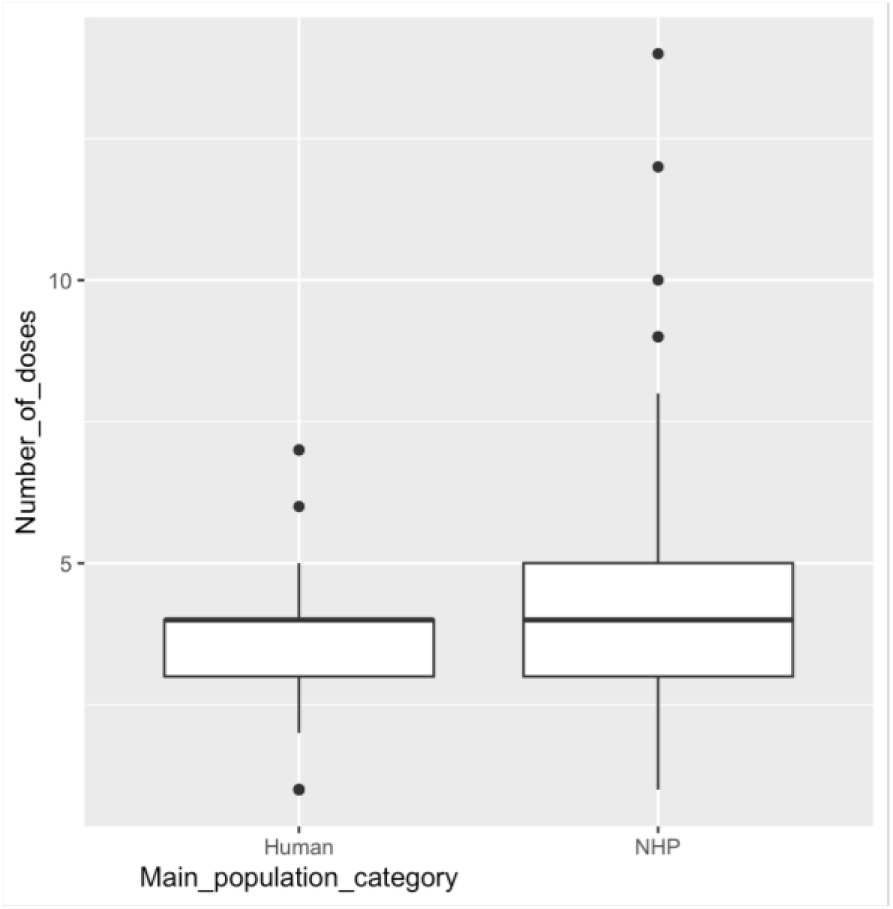
Number of doses in human and NHP studies.

**Figure 7:**
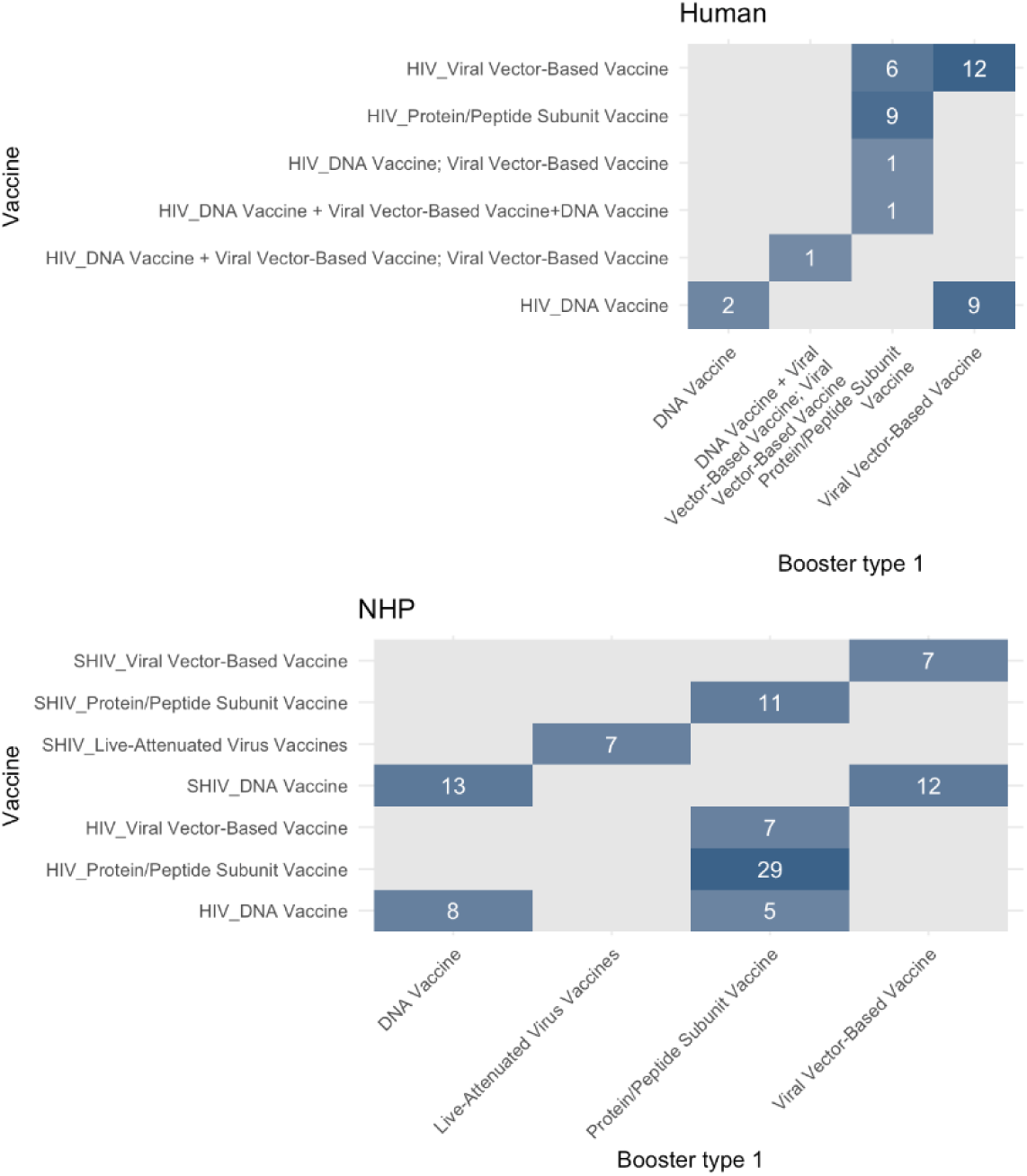
Heatmap of vaccine-booster combinations in humans and NHPs.

All included studies used between-subject controls. All included human studies compared vaccine administration with placebo. In NHP studies, the most common control conditions were Vehicle (k=69); Untreated (k=64); and Placebo (k=22). Other NHP studies used combinations of different control conditions (k=14), or a Sham control (k=1).

Overall, study durations from vaccination to outcome measurement ranged from 1 to 2099 days (median: 449.5). For NHP studies this entire range was covered (median: 469 days), for human studies study durations ranged from 121 to 1821 days (median: 365 days). Human studies were mainly performed in both low-risk (k=28) and high-risk (k=10) populations. NHP studies implemented challenges (described in k=170 of the included NHP studies).

We extracted data from the main efficacy outcomes for each vaccine-control comparison within each included study. In human studies, infectivity was the most frequently reported (c=104 out of c=114 human comparisons, versus c=113 out of c=490 NHP comparisons), while viral presence (mainly viral load) was the most frequently reported in NHP studies (c=336 comparisons, versus c=7 human comparisons). Mortality could only be extracted from c=40 of the NHP comparisons.

In human subjects, high risk of bias was mainly related to differences in baseline characteristics (k=11) and other sources of bias (k=10). Low risk of bias was most frequently observed for blinding of the intervention throughout the trial (k=44) and complete reporting of outcome data (k=36). Overall, unclear risk of bias scores were most common (202 unclear scores out of 414 total scored elements in human studies). In NHPs, high risk of bias was mainly related to incomplete outcome data (k=48), and other sources of bias (k=25). Low risk of bias was only frequently observed for complete reporting of outcome data (k=92). In NHP studies, unclear risk of bias scores were even more common (1480 unclear scores out of 1690 total scored elements in NHP studies). The risk of bias per scored element is shown in Figure 8.

**Figure 8:**
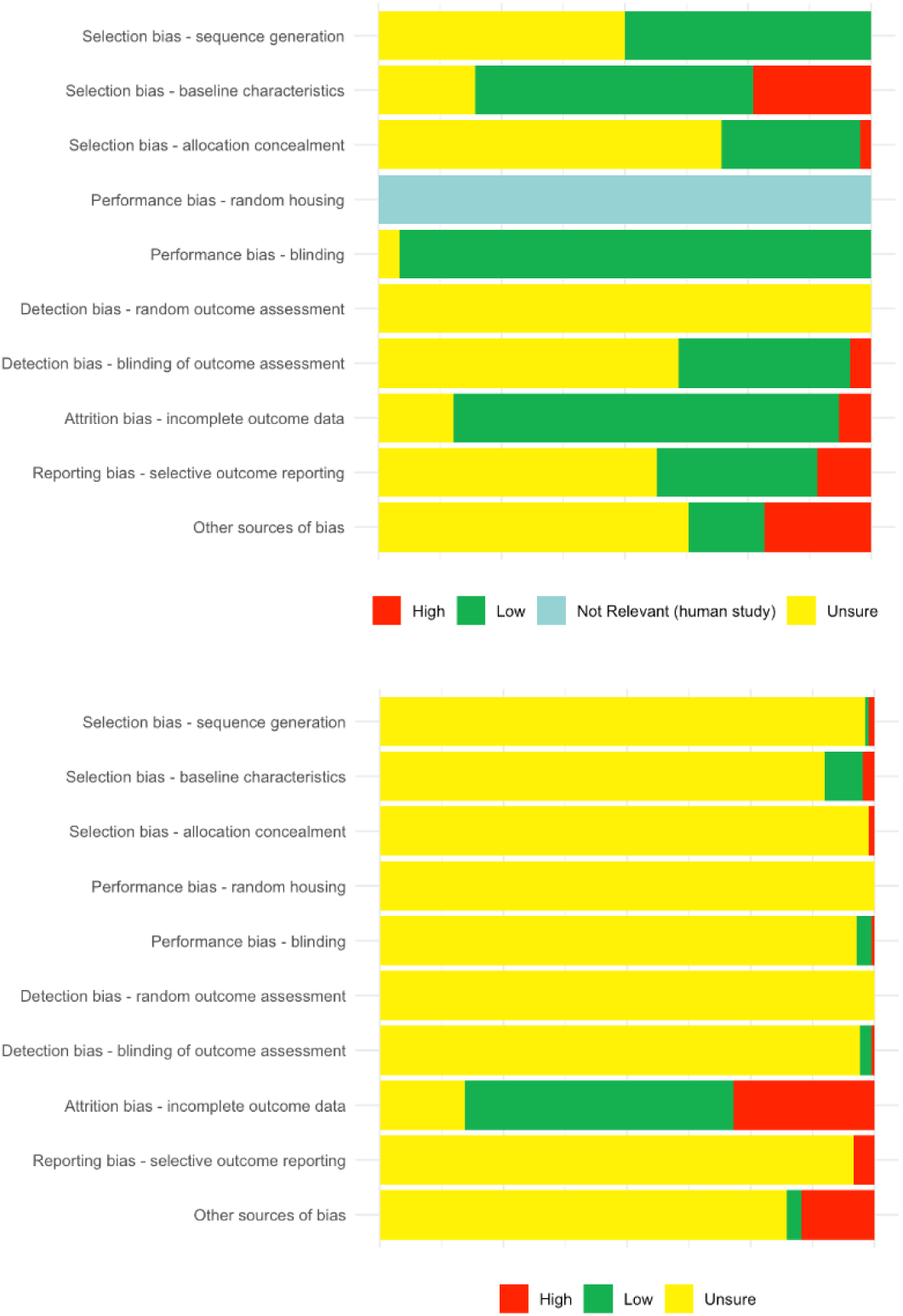
Risk of Bias in human (top graph) and NHP (bottom graph) studies.

An explorative overall random effects meta-analysis of the Risk Ratio (RR) of infection/ disease/ death in vaccinated subjects versus controls, sub-grouped by NHP vs. human subjects, showed a small decrease in overall risk after vaccination (Overall RR: 0.92; 95% confidence interval (CI): 0.89-0.95; z =-4.43; p<0.0001, Heterogeneity: I² = 15.8%), without a significant difference between animals and humans (Q=1.75; df=1; p=0.19). RRs per included study with the study 95%-CI are shown in Figure 9. A sensitivity analysis restricted to studies that had at least one observation in all cells (i.e. infected and non-infected subjects in both vaccinated and control groups) came to the same conclusion (Overall RR: 0.93; 95% confidence interval (CI): 0.89-0.96; z =-4.11; p<0.0001, Heterogeneity: I² = 10.2%).

**Figure 9:**
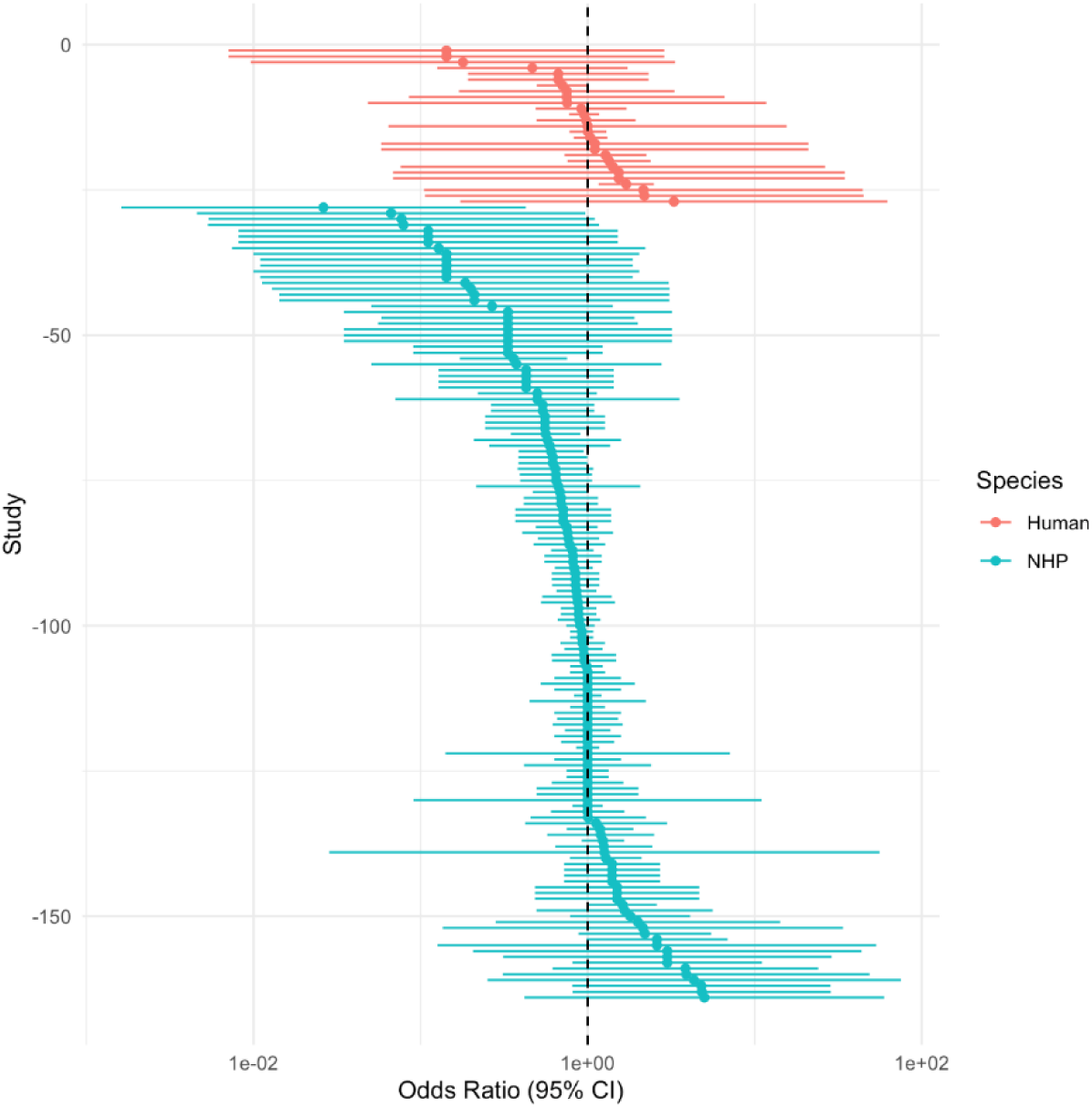
Forest plot of the outcomes of the included studies with their 95% confidence intervals.

The trim and fill analysis (Figure 10) showed the presence of small study effects. The adjusted overall effect size after adding 17 studies was 0.93; 95% CI: 0.89-0.96; z=-4.09; p< 0.0001).

**Figure 10:**
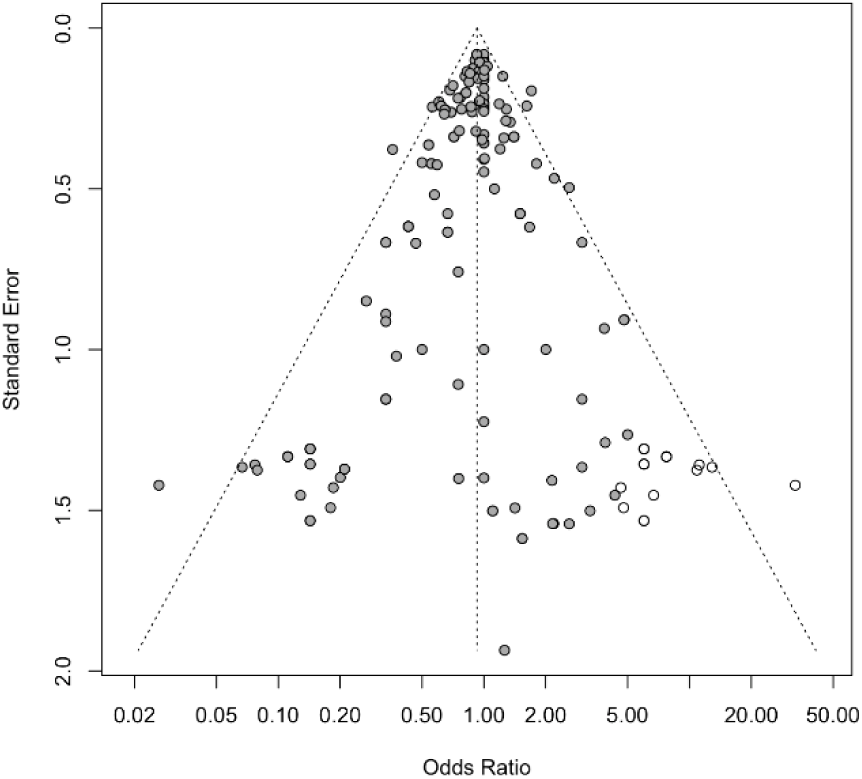
Funnel plot with trim & fill analysis. Filled circles: data from the included studies. Open circles: fictive studies added in the trim & fill analysis.

The percentages of NHP and human subjects infected in vaccinated and control groups per study is shown in Figure 11. While not formally analyzed, NHP studies clearly have higher overall infection rates than human studies (probably due to challenges).

**Figure 11:**
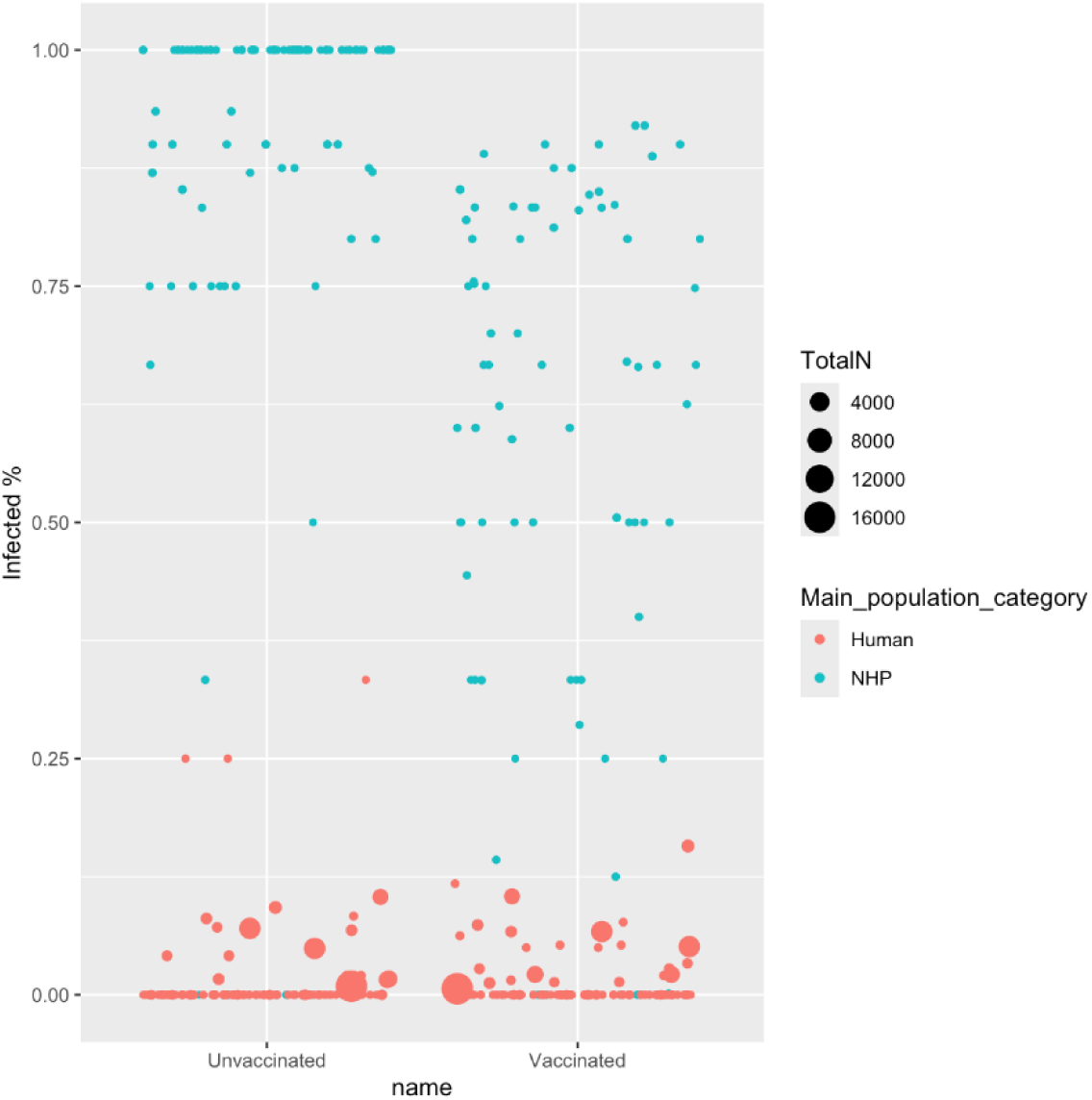
Bubble plot of infection rates.

We originally intended to explore potential CoPs and extracted proxy outcomes for efficacy. A word cloud of these extractions is provided in Figure 12. The most commonly mentioned proxy outcomes were antibodies (k=147), cells (k=106), elispot (k=78) and CD4 (k=77). Again, these values are underestimates because we did not correct for typographical errors and variant spellings.

**Figure 12:**
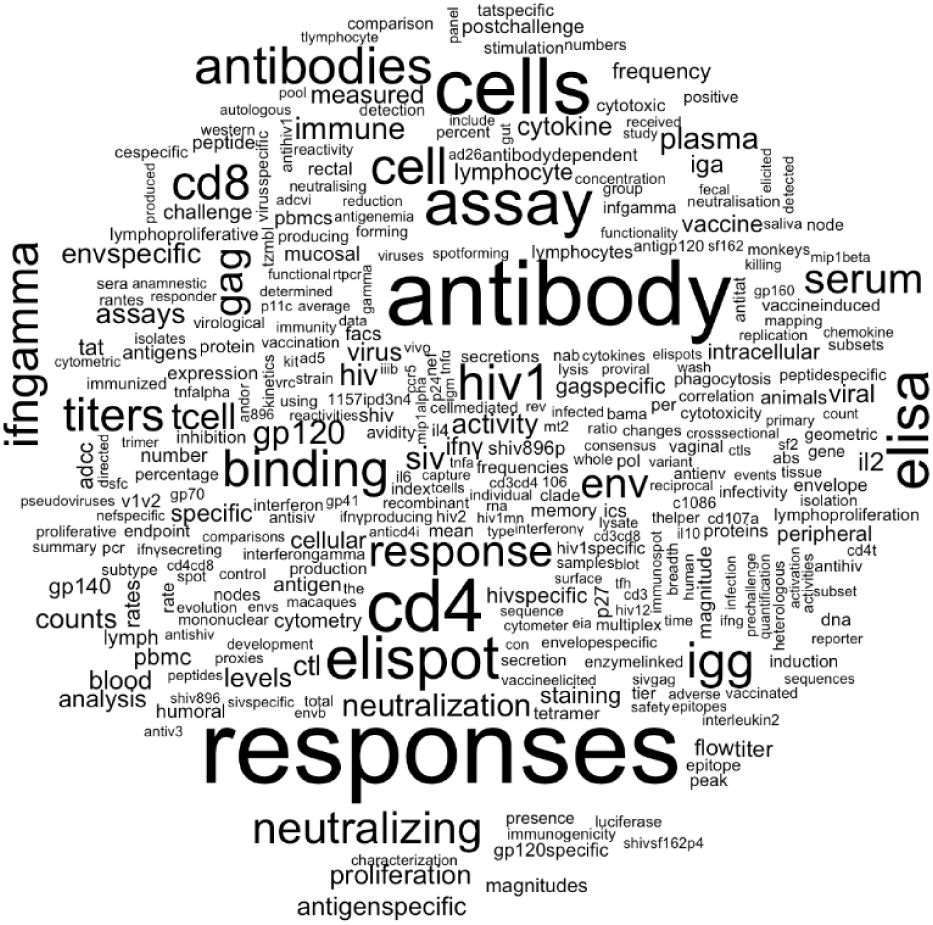
Word cloud of mentioned proxy outcomes for efficacy.

To aid scientists working on HIV, we have made our extracted data available in a spreadsheet. While the data were not collected with the intention of creating a clean and easily searchable database, this spreadsheet can be used to retrieve all relevant studies on any specific topic up to 25 December 2023 by simple text term searches and/ or filtering the values for variables of interest.

## Discussion

This SR summarizes the findings of 216 scientific papers describing HIV efficacy studies in humans and NHPs. For the 46 papers describing human studies, 40 750 volunteers were exposed to experimental HIV vaccines; for the 170 papers describing NHP studies, 3 326 NHPs were exposed. These 44 076 experimentally vaccinated subjects have not resulted in a regulatory approved vaccine for humans, nor in a reliable CoP being defined. Relevant differences were observed in the designs of NHP and human studies for the number of subjects per group, the study duration, the number and type of vaccines administered, the control conditions, the level of reporting experimental detail (resulting in differences in risk of bias estimations) and the level of infection (presumably related to challenges in NHP studies). For at least group size and risk of bias, similar differences have been observed and reported between animal and human studies in other fields(62–64). In spite of these differences in designs, overall the efficacy results for NHP and human HIV vaccine efficacy studies show a high degree of overlap, probably because “nothing really works” and confidence intervals are large. At this stage, limited animal-to-human translation can therefore not be held responsible for not yet finding an effective preventive HIV vaccine.

### Strengths and limitations

This is one of the first SRs quantitatively comparing results from animal and human vaccine studies, and as far as the authors are aware, the very first addressing the efficacy of HIV vaccines. The comprehensive search strategy ensures capturing virtually all information meeting the inclusion criteria up to the search date (25-DEC-2023). The robust methods implemented throughout, with each reference being evaluated by two reviewers in all stages, ensure that our conclusions are as reliable as possible. Providing open access to our extracted data in a spreadsheet can aid scientists in the field to retrieve relevant studies more easily.

While we had to adapt our plans compared to our protocol because of the number of retrieved references exceeding our expectations and capacity, our mainly minor protocol deviations were made in a manner that should not result in review bias. The main disadvantage results from not being able to contact authors of included studies for supplying missing data and exclusion of SIV vaccines. Although author response rates in requests from systematic reviewers vary (65, 66), adding additional data would have increased the informative value of both our data spreadsheet and our analyses.

Comparably, without analyses of SIV vaccines in NHPs, we cannot assess the value of SIV-NHP models. However, not analysing the proxy outcomes for vaccine efficacy had no relevant consequences, as potential CoPs cannot be defined before reliable positive efficacy results become available (23).

The main limitation of this review is that it was focused solely on efficacy of preventive vaccines, reflecting the inherent compromise between specificity and comprehensiveness that arises from applying strict inclusion and exclusion criteria in systematic reviews(67, 68). While necessary for reliability and reproducibility, these criteria restrict both the scope and the depth of the SR. Anything not meeting the criteria is excluded and cannot be analyzed, but using a wide scope can also restrict the viable depth of the analyses. For this manuscript, we provide a reliable overview of the current status of HIV vaccine efficacy research in NHPs and humans. However, similar SRs ought to be performed addressing vaccine safety, therapeutic vaccination and passive immunization, which have currently only been reviewed narratively (15, 22). Besides, more detailed and in-depth SRs of e.g. specific models, specific epitopes or specific vaccine-boost strategies might shed more light on their value.

### Hurdles to HIV vaccine development

Loosely based on the work described above, we identified three potential hurdles in HIV vaccine development which we discuss below: first, too much variation in tested strategies; second, suboptimal experimental designs; and third, misleading communication.

To start with variation, it is in human nature to keep doing something if it works, and to explore alternative strategies if it does not. For HIV vaccine development, scientists initially tested multiple approaches that had been effective for other pathogens and later moved on to exploring more novel ideas. Without successes, the approaches have continued to diverge over time, resulting in multiple strategies being explored without sufficient replications. Of course, studying alternative vaccination strategies would be easier with a reliable CoP, but none has been identified for HIV (23). The most informative findings about potential CoPs stem from the only clinical trial that has demonstrated some vaccine efficacy to date (albeit low and short): the RV144 trial (69). The vaccine efficacy shown in this trial was unexpected for multiple reasons: the vector used was less immunogenic than others, the booster was not effective in other trials, and the heterosexual Thai population was not at high risk of getting infected (70). Subsequent analyses of the data and available samples mainly used a case-control approach, and comparisons with negative findings from other trials(70, 71). Within RV144, non-neutralizing antibodies to specific viral epitopes, polyfunctional CD4+ T-cell responses to envelope peptides and specific host polymorphisms were correlated with vaccine efficacy, while high serum IgA antibodies to envelope epitopes were inversely correlated (possibly because of hindering antibody-dependent cellular cytotoxicity). Vaccine development incorporated these putative CoPs, but also kept adapting approaches based on broad neutralizing antibodies and more classical approaches(7, 11, 16, 17).

By now, a multitude of different vaccine regimens have been tested in NHPs and humans. Most of these regimens were safe, and many were also immunogenic. However, immunogenicity was not generally paired with adequate levels of protection (72). Overall, this multipronged approach may hinder the determination of immune CoPs (73). To complicate matters even further, CoPs may well vary for different routes of infection. In spite of relative inefficiency and while there are large regional differences in transmission routes, overall, most HIV transmission is currently via heterosexual contact (71). However, local immune responses at the most common sites of infection (male and female genital mucosa) have hardly been measured for several reasons, most importantly the increased risk of infection with impaired barrier function (21, 23, 25, 71). Although challenging, it may be sensible to focus further research more in this direction.

While HIV’s very effective manner of evading the immune system might still require novel solutions for candidate vaccine regimes, the current width of testing various strategies without much replication of studies does not allow for meaningful data syntheses. At this stage, in-depth SRs of e.g. different vaccine types, induction of broad neutralizing antibodies, induction of non-neutralizing antibodies, induction of cellular responses, induction of the innate immune response, mosaic vaccines, various adjuvants, etc. have been tested in only a few studies and with substantial variation in designs (Figures 5 & 7). Thus, in-depth SRs of these strategies will probably not be informative yet.

To continue with suboptimal experimental designs; two main concerns arose: control conditions and statistical power. The included between-subject control conditions mainly comprised placebo, vehicle and untreated. Although we only extracted data from the *most* comparable control group(s), these results confirm a preceding notion that controls in HIV studies may be oversimplified (23). Vaccine formulations comprise multiple compounds besides the antigen, and mainly the adjuvants could boost the recipients’ immune responses in non-specific manners. Also, the presence of foreign DNA, RNA or proteins, regardless of the antigenic epitope, could have such effects. Thus, the use of proper control formulations containing all non-vaccine compounds, besides non-viral (sequence-scrambled) DNA/ RNA/ proteins, all in equivalent amounts, should be advocated.

Statistical power is an important part of any experimental design, and a general problem in preclinical animal research, where underpowered studies are relatively common (74). With low numbers of subjects tested in several of the animal and human studies (Figure 4), the HIV vaccine development field is no exception. To this end, it is concerning that adaptive clinical trial designs are advocated(21, 70). Caution against implementation of adaptive trial designs is warranted, as they can reduce the statistical power and make outcomes less reliable and informative.

To finish with scientific communication, misleading phrasing may be an issue. Many of the included publications and relevant reviews were written using optimistic phrasing such as “shows that an effective vaccine is within reach” (16, 18, 25), also in combination with negative/ neutral findings. In line with this optimism, publications have listed many ideas to improve future vaccine design and vaccination schedules, comprising novel epitopes, combinations and / or exchange of epitopes (mosaic), combined elicitation of humoral and cellular immune responses, innovative use of boosters, inventive prime-booster schedules, increasing the innate immune response, masking of immunodominant regions, structural stabilization of the virus particles, strategies comprising non-neutralizing antibodies targeting FCγ receptors, etc. (11, 12, 16, 19, 75). However, as described above, the field is already following highly varied approaches, and optimistically adding to the variation may not be fruitful.

Besides optimism, oversimplification seems common in the field; the lack of full understanding is hardly addressed. Limits in understanding are clear from the observations that certain experimental vaccine regimes, that were thought to be protective and tested in clinical trials, increased the risk of infection instead (6, 7, 23, 25). While vaccine development for other pathogens has largely been based on CoPs, many of the there-useful correlates are not mechanistic (7, 23, 76). For other pathogens (such as HIV), measurement of (neutralizing) antibodies is simply not sufficient (7, 76). If neutralizing antibody responses against a pathogen are present, they may not neutralize new variants, but vaccinees can still be protected from severe disease in spite of getting infected. While the more informative mechanistical CoPs are sought after, it is generally ignored that they may well vary with age, route of transmission/ measured compartment, innate immune system, comorbidities, history of other infections, vaccinees’ microbiomes and many other factors (7, 17, 23, 76). Simplification of a system as complex as the immune response to pathogens may be efficient in creating vaccines for more straightforward pathogens. However, it has not proven successful in developing an HIV vaccine. In line with this increasing knowledge, continued optimistic phrasing of negative/ neutral study results can be misleading and counterproductive.

### The value of NHP models in HIV vaccine research

Our main review question addressed the concordance of candidate HIV vaccine efficacy in non-human primates and humans. While the overall concordance was better than the review team anticipated, we are not confident in the predictive value of NHP models for human trial outcomes. Most scientists consider NHP models to be informative (25, 77), but this is mainly an assumption based on qualitative comparisons of NHP and human pathology. As described in the introduction, quantitative comparisons have thus far been lacking, the here-presented one is a first. However, we pooled the included studies regardless of study design and vaccine regime, while their variation between the included HIV trials is substantial. When *individual* vaccine regimens are compared, the same regimen can have either similar or opposite results in NHPs and humans (23). An illustrative example is the observation that the non-neutralizing antibodies that correlated with protection in the RV144 trial are not commonly observed in preclinical studies (73). The current review did identify problems in the alignment of trial design and in the reporting of experimental details. In the authors’ opinions, it does not contradict a previous statement that NHP trials are not useful as gatekeepers for human clinical trials (23).

NHP models and humans differ in many relevant aspects, such as cellular and humoral immune responses, protective epitopes, protective antibody concentrations, barrier functions at the various possible sites of infection and spontaneous infection (16, 17, 21, 25, 26). Our knowledge of the comparability of potentially protective immune responses between species is limited (70). In the absence of quantitative comparisons and a clear definition of what constitutes an acceptable predictive value, a review of ideas on how to improve animal-to-human predictability for vaccine efficacy might be considered premature (36).

Besides the biological differences, we observed relevant differences in the design and reporting of NHP and human studies (discussed above). Others noted that NHP models use challenges that are more aligned and homogeneous with the vaccines than can be expected in humans (25). In spite of a 2010 workshop of leading HIV-1 scientists advocating parallel and directly comparable NHP and human trials (21), we only included one paper describing both NHP and human findings using a comparable regimen (60). While animal and human studies can be published separately in spite of being aligned, based on what we read, this practice does not seem common (12, 23). Separately published yet aligned NHP-human trials are difficult to retrieve without additional effort, but it could be of interest to quantitatively synthesize the level of NHP-human agreement over multiple studies of comparable interventions.

### Future perspectives

To conclude, while *in vitro* studies of animal and human immune systems remain challenging (17, 18) and preclinical studies can result in mechanistic findings (6, 18), translation to humans is far from certain, making the value of studies using NHP models questionable. With or without preclinical NHP models, developing an effective HIV vaccine remains a worthwhile endeavor. The global incidence of HIV/AIDS reached 1.65 million in 2021, which translates to over 4500 new cases each day. While future *incidence* could either increase or decline (78, 79), with patients aging, the HIV burden is certainly expected to increase at least in the near future. Health economic modelling has convincingly shown that HIV vaccines can be cost-effective in many different populations (10). From the economical perspective, moderate effectiveness and limited durability of protection could be compensated by frequent boosting, but the economic models did not incorporate further costs of vaccine development. Besides, herd immunity limiting transmission, which was incorporated in some of the models, is based on good vaccine coverage.

While investing further funds into developing an HIV vaccine thus remains worthwhile, future coverage could be problematic, as high efficacy is a crucial factor in vaccine acceptability (5, 80). Besides, four decades of research and development without a safe and effective vaccine being found show that it is incredibly hard to develop an effective HIV vaccine. While we partially start to understand the multitude of reasons why HIV is so complicated to protect against (15), we may need to at least consider the possibility that developing an effective vaccine may be not possible.

## Statements

### Ethics review

No ethical review was conducted for this systematic review.

### Data availability

All extracted data are available from(https://osf.io/zkmcp). Source data are available in the original publications for which references are provided per review phase in the data files.

### Funding

A grant specific for this SR was provided by the USA-based nonprofit animal rights organization “People for the Ethical Treatment of Animals” (PETA).

Overall, the team members performing this review, which relates to the expected benefit and predictive value of animal experiments, are not prejudiced towards or against the use of animal experiments; we have a combination of team members with varying opinions. Some are convinced that there is no adequate evidence for the benefit from animal experiments, which means that we should not be doing animal experiments, some are unconvinced either way, which means that we should at least further analyse the available data before planning new animal experiments, and some believe that animal experiments can be informative for human health, as long as they are performed in a scientifically sound manner.

The review was performed as rigorous and as blind to the outcomes as realistically possible, to prevent reviewers’ opinions affecting the results. The authors did not expect to find the here-reported degree of overlap between NHP and human studies.

## Acknowledgements

The authors gratefully acknowledge Felix Weijdema and Alice Tillema for valuable advice on the search strategy; Laurens De Bruin for spontaneous assistance in data management; Rens van de Schoot for advice on AI-assisted screening and ASReview; and Jeffrey Bajramovic for general background information and advice on which data to extract and pool.

Any errors in this manuscript remain the responsibility of the authors.

