## Appendix A for "The efficacy of candidate HIV vaccines in non-human primates and humans: a systematic review"

**Search Report**

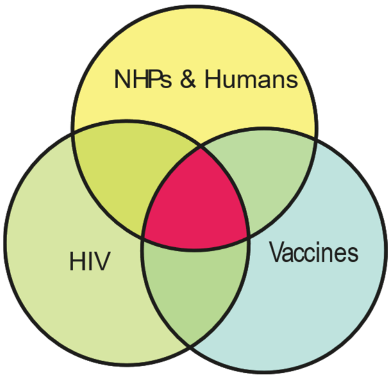

Separate searches were performed for NHP and human studies, on the same date (two sequential days for the two databases). The studies retrieved by both searches were taken out of the human batch via NOT combinations to prevent unnecessary screening of the same studies twice. The thus-created batches were screened separately, but NHP and human studies meeting the other criteria were included regardless of batch. The figure depicts the overall structure, the part in reddish is what has been screened.

| String/ Block | | #hits* |
|---|---|---|
| **PubMed** | | **25 DEC 23** |
| HIV | AIDS virus [MeSH] OR HIV [MeSH] OR HIV Infections[MeSH] OR "Human immuno deficiency virus"[TiAb] OR "Human immunodeficiency virus" [TiAb] OR "Human immunodeficiency viruses" [TiAb] OR "Lymphadenopathy-associated virus" [TiAb] OR "immunodeficiency associated virus" [TiAb] OR "lymphadenopathy associated retrovirus" [TiAb] OR "lymphadenopathy associated retroviruses" [TiAb] OR "Immunodeficiency Virus, Human" [tiab] OR "Immunodeficiency Viruses, Human"[tiab] OR "Virus, Human Immunodeficiency"[tiab] OR "Viruses, Human Immunodeficiency"[TiAb] OR "Human T-Cell Lymphotropic Virus Type III"[tiab] OR "Human T-Cell Leukemia Virus Type III"[tiab] OR "Human T-Lymphotropic Virus Type III"[tiab] OR "Acquired Immune Deficiency Syndrome"[tiab] OR "Acquired Immunodeficiency Syndrome"[tiab] OR “HTLV-III”[tiab] OR "Human T-Lymphotropic Virus Type IV"[tiab] OR "HTLV-IV"[tiab] OR "SBL-6669"[tiab] OR "Immunodeficiency Virus Type 1, Human"[tiab] | 384 324 |
| Vaccines | "aids vaccines"[MeSH] OR vaccin*[tiab] OR immunis*[tiab] OR immuniz*[tiab] | 515 838 |
| NHPs* | “catarrhini”[MeSH:noexp] OR “cercopithecidae”[MeSH] OR “gorilla gorilla”[MeSH] OR “haplorhini”[MeSH:noexp] OR“hominidae”[MeSH:noexp] OR “hylobatidae”[MeSH] OR “pan paniscus”[MeSH] OR “pan troglodytes”[MeSH] OR “platyrrhini”[MeSH] OR“pongo”[MeSH] OR “primates”[MeSH:noexp] OR “strepsirhini”[MeSH] OR“tarsii”[MeSH] OR “allenopithecus”[tiab] OR “allocebus”[tiab] OR“alouatta”[tiab] OR “alouattinae”[tiab] OR “angwantibo*”[tiab] OR“anthropoid”[tiab] OR “anthropoidea”[tiab] OR “anthropoids”[tiab]OR “aotes”[tiab] OR “aotidae”[tiab] OR “aotinae”[tiab] OR“aotus”[tiab] OR “ape”[tiab] OR “apes”[tiab] OR “arctocebus”[tiab]OR “ateles”[tiab] OR “atelidae”[tiab] OR “atelinae”[tiab] OR“avahi”[tiab] OR “aye-aye*”[tiab] OR “baboon”[tiab] OR“baboons”[tiab] OR “bonobo”[tiab] OR “bonobos”[tiab] OR“brachyteles”[tiab] OR “bushbabies”[tiab] OR “bushbaby”[tiab] OR "bush babies"[tiab] OR "bush baby"[tiab] OR“cacajao”[tiab] OR “callibella”[tiab] OR “callicebinae”[tiab] OR“callicebus”[tiab] OR “callimico”[tiab] OR “callithrichid*”[tiab] OR“callithrichinae”[tiab] OR “callithrix”[tiab] OR “callitrichid”[tiab] OR“callitrichidae”[tiab] OR “callitrichide”[tiab] OR “callitrichids”[tiab]OR “callitrichinae”[tiab] OR “capuchin”[tiab] OR “capuchins”[tiab]OR “carlito | 266 528 |

| | |
|---|---|
| | syrichta”[tiab] OR “catarhine*”[tiab] OR “catarhini”[tiab]OR “catarrhina”[tiab] OR “catarrhine*”[tiab] OR “catarrhini”[tiab]OR “cebid”[tiab] OR “cebidae”[tiab] OR “cebids”[tiab] OR“cebinae”[tiab] OR “ceboidea”[tiab] OR “cebuella”[tiab] OR“cebus”[tiab] OR “cephalopachus”[tiab] OR “cercocebus”[tiab] OR“cercopithecid*”[tiab] OR “cercopithecinae”[tiab] OR“cercopithecine*”[tiab] OR “cercopithecini”[tiab] OR“cercopithecoid”[tiab] OR “cercopithecoidea”[tiab] OR“cercopithecoids”[tiab] OR “cercopithecus”[tiab] OR“cheirogaleidae”[tiab] OR “cheirogaleus”[tiab] OR“cheracebus”[tiab] OR “chimp”[tiab] OR “chimpanzee”[tiab] OR“chimpanzees”[tiab] OR “chimps”[tiab] OR “chiromyiformes”[tiab]OR “chiropotes”[tiab] OR “chlorocebus”[tiab] OR “colobidae”[tiab]OR “colobinae”[tiab] OR “colobine*”[tiab] OR “colobini”[tiab] OR“colobus*”[tiab] OR “cynomolgus”[tiab] OR “daubentonia”[tiab] OR“daubentoniidae”[tiab] OR “douc”[tiab] OR “doucs”[tiab] OR“erythrocebus”[tiab] OR “eulemur”[tiab] OR “euoticus”[tiab] OR“euprimate*”[tiab] OR “galagid*”[tiab] OR “galago”[tiab] OR“galagoides”[tiab] OR “galagonidae”[tiab] OR “galagos”[tiab] OR“gelada”[tiab] OR “geladas”[tiab] OR “gibbon”[tiab] OR“gibbons”[tiab] OR “gorilla”[tiab] OR “gorillas”[tiab] OR“grivet”[tiab] OR “grivets”[tiab] OR “guenon*”[tiab] OR“guereza*”[tiab] OR “hapalemur”[tiab] OR “haplorhine*”[tiab] OR“haplorhini”[tiab] OR “haplorrhine*”[tiab] OR “haplorrhini”[tiab] OR“hominid*”[tiab] OR “hominin”[tiab] OR “homininae”[tiab] OR“hominine”[tiab] OR “hominines”[tiab] OR “hominini”[tiab] OR“hominins”[tiab] OR “hominoidea”[tiab] OR “hoolock”[tiab] OR“howler*”[tiab] OR “hylobates”[tiab] OR “hylobatidae”[tiab] OR“indri”[tiab] OR “indridae”[tiab] OR “indriid*”[tiab] OR “indris”[tiab]OR “kipunji*”[tiab] OR “lagothrix”[tiab] OR “langur”[tiab] OR“langurs”[tiab] OR “lemur”[tiab] OR “lemurid*”[tiab] OR“lemuriform”[tiab] OR “lemuriformes”[tiab] OR “lemuriforms”[tiab]OR “lemurinae”[tiab] OR “lemuroidea”[tiab] OR “lemurs”[tiab] OR“leontideus”[tiab] OR “leontocebus”[tiab] OR “leontopithecus”[tiab]OR “lepilemur”[tiab] OR “lepilemurid*”[tiab] OR “lesula*”[tiab] OR“lophocebus”[tiab] OR “loriform”[tiab] OR “loriformes”[tiab] OR“lorinae”[tiab] OR “loris”[tiab] OR “lorises”[tiab] OR “lorisid*”[tiab]OR “lorisiform*”[tiab] OR “lorisinae”[tiab] OR “lorisoid*”[tiab] OR“lutung”[tiab] OR “lutungs”[tiab] OR “macaca”[tiab] OR“macaque's”[tiab] OR “macaque”[tiab] OR “macaques”[tiab] OR“malbrouck*”[tiab] OR “mandrill”[tiab] OR “mandrills”[tiab] OR “mandrillus”[tiab] OR “mangabey*”[tiab] OR “marmoset”[tiab] OR “marmosets”[tiab] OR “mico argentatus”[tiab] OR “mico chrysoleucos”[tiab] OR “mico chrysoleucus”[tiab] OR “mico emiliae”[tiab] OR “mico humilis”[tiab] OR “mico marcai”[tiab] OR “mico melanurus”[tiab] OR “mico rondoni”[tiab] OR “microcebus”[tiab] OR “miopithecus”[tiab] OR “mirza coquereli”[tiab] OR “mirza zaza”[tiab] OR “monkey”[tiab] OR “monkeys”[tiab] OR “muriqui*”[tiab] OR “nasalis larvatus”[tiab] OR “nomascus”[tiab] OR “nycticebus”[tiab] OR “oedipomidas”[tiab] OR “orang utan*”[tiab] OR “orang-utan*”[tiab] OR “orangutan*”[tiab] OR “oreonax”[tiab] OR “otolemur”[tiab] OR “pan paniscus”[tiab] OR “pan troglodytes”[tiab] OR “panin”[tiab] OR “panina”[tiab] OR “panins”[tiab] OR “papio”[tiab] OR “papionini”[tiab] OR “paragalago”[tiab] OR “perodicticinae”[tiab] OR “perodicticus”[tiab] OR “phaner”[tiab] OR “piliocolobus”[tiab] OR “pithecia”[tiab] OR “pithecidae”[tiab] OR “pitheciid*”[tiab] OR “pitheciin*”[tiab] OR “pithecin*”[tiab] OR “platyrhine*”[tiab] OR “platyrhini”[tiab] OR “platyrrhina”[tiab] OR “platyrrhine*”[tiab] OR “platyrrhini”[tiab] OR “plecturocebus”[tiab] OR “pongid*”[tiab] OR “ponginae”[tiab] OR “pongo”[tiab] OR “potto”[tiab] OR “pottos”[tiab] OR “presbytini”[tiab] OR “presbytis”[tiab] OR “primate”[tiab] OR “primates”[tiab] OR “procolobus”[tiab] OR “prolemur”[tiab] OR “propithecus”[tiab] OR “prosimian*”[tiab] OR “prosimii”[tiab] OR “pseudopotto”[tiab] OR “pygathrix”[tiab] OR “rhinopithecus”[tiab] OR “rungwecebus”[tiab] OR “saguinus”[tiab] OR “saimiri”[tiab] OR “saimiriinae”[tiab] OR “sapajus”[tiab] OR “sciurocheirus”[tiab] OR “semnopithecus”[tiab] OR “siamang”[tiab] OR “siamangs”[tiab] OR “sifaka”[tiab] OR “sifakas”[tiab] OR “simians”[tiab] OR |

| | | |
|---|---|---|
| | “simias”[tiab] OR “simiiform*”[tiab] OR “strepsir*”[tiab] OR “surili*”[tiab] OR “symphalangus”[tiab] OR “talapoin*”[tiab] OR “tamarin”[tiab] OR “tamarins”[tiab] OR “tamarinus”[tiab] OR “tarsier”[tiab] OR “tarsiers”[tiab] OR “tarsiid*”[tiab] OR “tarsiiform*”[tiab] OR “tarsius”[tiab] OR “theropithecus”[tiab] OR “trachypithecus”[tiab] OR “uacari*”[tiab] OR “uakari”[tiab] OR “uakaris”[tiab] OR “varecia”[tiab] OR “vervet*”[tiab] | |
| Humans** | “clinical study”[pt] OR “randomized controlled trial”[pt] OR “controlled clinical trial”[pt] OR humans[MeSH] OR “Clinical Trials as Topic”[MeSH] OR “intervention study”[tiab] OR “first-in-man”[tiab] OR trial[tiab] OR human[tiab] OR humans[tiab] | 22 539 370 |
| Combined results for PubMed (25-DEC-2023):<br>20 454 hits overall<br>2 709 potentially relevant mainly NHP references<br>20 169 potentially relevant mainly human references (17 745 of which not in the NHP batch)<br>2 424 of which are present in both sets | | |
| **Embase** | | **26 DEC 23** |
| HIV | Exp “Human immunodeficiency virus”/ OR “Human immunodeficiency virus infection”/ OR “acute HIV infection”/ OR “Human immunodeficiency virus 1 infection”/ OR “Human immunodeficiency virus 2 infection”/ OR (“Human immuno deficiency virus” OR “Human immunodeficiency virus” OR “Human immunodeficiency viruses” OR “Lymphadenopathy-associated virus” OR “immunodeficiency associated virus” OR “lymphadenopathy associated retrovirus” OR “lymphadenopathy associated retroviruses” OR “Immunodeficiency Virus, Human” OR “Immunodeficiency Viruses, Human” OR “Virus, Human Immunodeficiency” OR “Viruses, Human Immunodeficiency” OR “Human T-Cell Lymphotropic Virus Type III” OR “Human T-Cell Leukemia Virus Type III” OR “Human T-Lymphotropic Virus Type III” OR “Acquired Immune Deficiency Syndrome” OR “Acquired Immunodeficiency Syndrome” OR “HTLV-III” OR “Human T-Lymphotropic Virus Type IV” OR “HTLV-IV” OR “SBL-6669” OR “Immunodeficiency Virus Type 1, Human”).ti,ab,kf. | 506 081 |
| Vaccines | Exp “Human immunodeficiency virus vaccine”/ OR (vaccin* OR immunis* OR immuniz*).ti,ab,kf. | 597 157 |
| NHPs* | Primate/ OR “Exp Primate model”/ OR Exp Prosimian/ OR haplorhini/ OR Exp tarsiiform/ OR simian/ OR Exp platyrrhini/ OR catarrhini/ OR Exp cercopithecidae/ OR ape/ OR Exp hylobatidae/ OR hominid/ OR Exp chimpanzee/ OR Exp gorilla/ OR Exp orangutan/ OR (allenopithecus OR allocebus OR alouatta OR alouattinae OR angwantibo* OR anthropoid OR anthropoidea OR anthropoids OR aotes OR aotidae OR aotinae OR aotus OR ape OR apes OR arctocebus OR ateles OR atelidae OR atelinae OR avahi OR “aye-aye*” OR baboon OR baboons OR bonobo OR bonobos OR brachyteles OR bushbabies OR bushbaby OR "bush babies" OR "bush baby" OR cacajao OR callibella OR callicebinae OR callicebus OR callimico OR callithrichid OR callithrichinae OR callithrix OR callitrichid OR callitrichidae OR callitrichide OR callitrichids OR callitrichinae OR capuchin OR capuchins OR “carlito syrichta” OR catarhine* OR catarhini OR catarrhina OR catarrhine* OR catarrhine OR cebid OR cebidae OR cebids OR cebinae OR ceboidea OR cebuella OR cebus OR cephalopachus OR cercocebus OR cercopithecid* OR cercopithecinae OR cercopithecine* OR cercopithecini OR cercopithecoid OR cercopithecoidea OR cercopithecoids OR cercopithecus OR cheirogaleidae OR cheirogaleus OR cheracebus OR chimp OR chimpanzee OR chimpanzees OR chimps OR chiromyiformes OR chiropotes OR chlorocebus OR colobidae OR colobinae OR colobine OR colobine OR colobus* OR cynomolgus OR daubentonia OR daubentoniidae OR douc OR doucs OR erythrocebus OR eulemur OR euoticus OR euprimate* OR galagid* OR galago OR galagoides OR galagonidae OR galagos OR gelada OR geladas OR gibbon OR gibbons OR gorilla OR gorillas OR grivet OR grivets OR guenon* OR guereza* OR hapalemur OR haplorhine* OR haplorhini OR haplorrhine* OR haplorrhini OR hominid* OR | 238 967 |

| | | |
|---|---|---|
| | hominin OR homininae OR hominine OR hominines OR hominini OR hominins OR hominoidea OR hoolock OR howler* OR hylobates OR hylobatidae OR indri OR indridae OR indriid* OR indris OR kipunji* OR lagothrix OR langur OR langurs OR lemur OR lemurid* OR lemuriform OR lemuriformes OR lemuriforms OR lemurinae OR lemuroidea OR lemurs OR leontideus OR leontocebus OR leontopithecus OR lepilemur OR lepilemurid* OR lesula* OR lophocebus OR loriform OR loriformes OR lorinae OR loris OR lorises OR lorisid* OR lorisiform* OR lorisinae OR lorisoid* OR lutung OR lutungs OR macaca OR “macaque's” OR macaque OR macaques OR malbrouck* OR mandrill OR mandrills OR mandrillus OR mangabey* OR marmoset OR marmosets OR “mico argentatus” OR “mico chrysoleucos” OR “mico chrysoleucus” OR “mico emiliae” OR “mico humilis” OR “mico marcai” OR “mico melanurus” OR “mico rondoni” OR microcebus OR miopithecus OR “mirza coquereli” OR “mirza zaza” OR monkey OR monkeys OR muriqui* OR “nasalis larvatus” OR nomascus OR nycticebus OR oedipomidas OR “orang utan*” OR “orang-utan*” OR orangutan* OR oreonax OR otolemur OR “pan paniscus” OR “pan troglodytes” OR panin OR panina OR panins OR papio OR papionini OR paragalago OR perodicticinae OR perodicticus OR phaner OR piliocolobus OR pithecia OR pithecidae OR pitheciid* OR pitheciin* OR pithecin* OR platyrhine* OR platyrhini OR platyrrhina OR platyrrhine* OR platyrrhini OR plecturocebus OR pongid* OR ponginae OR pongo OR potto OR pottos OR presbytini OR presbytis OR primate OR primates OR procolobus OR prolemur OR propithecus OR prosimian* OR prosimii OR pseudopotto OR pygathrix OR rhinopithecus OR rungwecebus OR saguinus OR saimiri OR saimiriinae OR sapajus OR sciurocheirus OR semnopithecus OR siamang OR siamangs OR sifaka OR sifakas OR simians OR simias OR simiiform* OR strepsir* OR surili* OR symphalangus OR talapoin* OR tamarin OR tamarins OR tamarinus OR tarsier OR tarsiers OR tarsiid* OR tarsiiform* OR tarsius OR theropithecus OR trachypithecus OR uacari* OR uakari OR uakaris OR varecia OR vervet*).ti,ab,kf. | |
| Humans** | Exp human/ OR “clinical study”/ OR Exp “clinical trial”/ OR (“intervention study” OR “first-in-man” OR trial OR human OR humans).ti,ab,kf. | 26 999 889 |
| Combined results for Embase (26-DEC-2023):<br><u>30170 hits overall</u><br>3 503 potentially relevant mainly NHP references<br>29 508 potentially relevant mainly human references (26 667 of which not in the NHP batch)<br>2 841 of which are present in both sets | | |

*NHP search string adapted from Cassidy et al. 2021 (dx.doi.org/10.1002/ajp.23287 ).
Adaptations to NHP filter for Pubmed: MH replaced by MeSH; "bush babies"[tiab] and "bush baby"[tiab] (with spacing) added; “mico chrysoleucus” (alternative spelling for Latin) added; pitheciinae[tiab] replaced by pitheciin*[tiab] and pithecinae[tiab] by pithecin*[tiab] to include alternate spellings.
Adaptations for Embase (from PubMed after above adaptations): MeSH terms replaced by Emtree terms following the respective hierarchy to exclude humans in the NHP string; syntax adapted.
**human search string adapted from Van de Wall et al. 2023; dx.doi.org/10.14573/altex.2208261
